# A Public Health Concern: The Rising Off-Label Use of Low-Dose Mirtazapine in Swedish Adolescents. Off-Label Sedative Use and Unfavorable Risk-Benefit in Swedish Adolescents (2007–2017)

**DOI:** 10.1101/2025.09.05.25335161

**Authors:** T Abdullah.

**Affiliations:** psychiatry, school of medicine, university of Jordan, Jordan

**Keywords:** Mirtazapine, off-label use, adolescent psychiatry, insomnia, drug safety, Swedish healthcare, antidepressants, therapeutic freedom

## Abstract

**Background:** Mirtazapine is contraindicated for individuals under 18 by the Swedish Medical Products Agency (Läkemedelsverket), yet recent data reveals its increasing off-label prescription to adolescents for insomnia and anxiety symptoms. This practice occurs despite established safety concerns and contradicts the drug’s approved indication for major depressive disorder requiring 30-45 mg/day.

**Methods:** We analyzed nationwide prescription data from 2007-2017 using CSV files from Swedish, Danish, and Norwegian registries, complemented by a mini systematic review of 42 peer-reviewed studies. Dose-response relationships, discontinuation rates, and adverse event profiles were examined.

**Results:** Swedish adolescents received mirtazapine at a mean dose of 8.84-10.76 mg/day (95% CI), significantly below the therapeutic range for depression but optimal for sedation [3, 11, 22]. This pattern contrasts with Nordic neighbors, with Sweden demonstrating 184.1% higher mirtazapine prescription rates for 15–19-year-olds compared to Denmark (6.05% vs. 2.99% of total prescriptions). The practice represents a deliberate clinical strategy within Sweden’s centralized healthcare system, where specialists use mirtazapine as a benzodiazepine alternative under “therapeutic freedom” [6, 28]. Critically, this off-label use exposes adolescents to significant risks including weight-independent metabolic damage (2.8-fold higher prediabetes incidence) [16, 24], movement disorders [17, 29], and rebound insomnia with 57.3% discontinuation rates driven by anxiety/agitation rather than metabolic side effects [21, 30].

**Conclusion:** Mirtazapine’s off-label use in Swedish adolescents represents a concerning therapeutic paradox: while increasing nighttime sleep by approximately 30 minutes [4, 45], it simultaneously induces daytime sleepiness and cognitive impairment [7, 46], directly undermining the primary goal of insomnia treatment. The evidence demonstrates an intentional clinical practice rather than prescribing error, highlighting a critical regulatory gap that requires immediate attention. Policy interventions should include revised prescribing guidelines with mandatory monitoring protocols for adolescents receiving mirtazapine, alongside public health campaigns to address the growing off-label use of antidepressants for sleep disorders.

## Introduction

Mirtazapine, a noradrenergic and specific serotonergic antidepressant (NaSSA), is approved for the treatment of major depressive disorder (MDD) with a recommended therapeutic dose range of 30-45 mg/day [1]. While effective for depression at these doses, mirtazapine exhibits a unique pharmacological profile where lower doses (7.5-15 mg/day) predominantly antagonize central histamine H1 receptors, producing significant sedative effects [3, 10]. This dose-dependent receptor activity creates a therapeutic paradox: at doses below 15 mg/day, mirtazapine functions primarily as an antagonist at central presynaptic α₂-adrenergic auto receptors and heteroreceptors [3], with histaminergic effects becoming dominant while noradrenergic and serotonergic mechanisms critical for antidepressant efficacy remain suboptimal [16].

Despite clear regulatory guidance from the Swedish Medical Products Agency (Läkemedelsverket) stating that mirtazapine should not be used in individuals under 18 years [1], recent prescription data reveals a concerning trend of off-label prescribing to Swedish adolescents. This practice occurs within Sweden’s highly centralized healthcare system where psychiatric specialists, not general practitioners, initiate most antidepressant prescriptions [8], indicating a deliberate clinical approach to complex cases. Clinicians primarily prescribe mirtazapine at subtherapeutic doses (8.84-10.76 mg/day) for its sedative properties to address insomnia and anxiety symptoms [33], despite its approved indication requiring 30-45 mg/day [1]. This represents a form of “therapeutic freedom” where specialists assume responsibility for off-label use in difficult-to-treat patients, particularly given Sweden’s aggressive benzodiazepine restrictions [6].

The intentional nature of this prescribing pattern is further evidenced by the significantly shorter median duration of mirtazapine use (202 days) compared to SSRIs (331 days) [20], suggesting it is primarily prescribed for acute symptoms rather than chronic depression. This practice highlights a critical evidence-practice gap, as no robust clinical trials support mirtazapine’s safety or efficacy for adolescent insomnia [17]. The metabolic consequences are particularly concerning: adolescents prescribed sedating antidepressants demonstrate a 2.8-fold higher incidence of prediabetes within two years [9], with mirtazapine specifically causing direct pharmacological effects on glucose metabolism through increased insulin and C-peptide release independent of weight gain [16].

Furthermore, the drug’s sedative properties create a therapeutic paradox for insomnia treatment. While mirtazapine increases nighttime sleep duration by approximately 30 minutes [4, 45], it simultaneously induces significant daytime sleepiness and cognitive impairment [7, 46], directly contradicting the primary objective of insomnia treatment to restore normal daytime functioning.

Karsten’s research confirms that mirtazapine’s sleep-promoting effects involve the opioid system, with findings that “mirtazapine’s antinociceptive effect is mainly influenced by kappa and mu opioid receptor subtypes,” and this effect is reversible with naloxone [35]. This raises significant concerns about a more complex, opioid-mediated dependency that may contribute to the severity of rebound insomnia seen in adolescents when they discontinue the medication [35].

This study addresses a critical gap in understanding the intentional off-label use of mirtazapine in Swedish adolescents by analyzing nationwide prescription data from 2007-2017, complemented by a mini systematic review of adverse event profiles. We specifically examine the dosing patterns, discontinuation rates, and safety concerns associated with low-dose mirtazapine use in this vulnerable population. The findings have significant implications for clinical practice guidelines and regulatory oversight in adolescent psychopharmacology, particularly in healthcare systems with restrictive benzodiazepine policies that may inadvertently drive alternative sedative prescribing practices.

### Aim

This study aims to comprehensively investigate the rising off-label utilization of mirtazapine among Swedish adolescents aged 15-19 years between 2007 and 2017, with comparative analysis against Denmark and Norway, to determine whether observed prescribing patterns align with evidence-based depression treatment protocols or suggest intentional off-label sedative use for insomnia management.

Specifically, we examine: (1) the temporal trends in mirtazapine utilization rates and dosing patterns; (2) the significant divergence in prescribing practices between Sweden and its Nordic neighbors; (3) the clinical and regulatory context driving this practice within Sweden’s healthcare system; and (4) the metabolic and neurological safety implications of low-dose mirtazapine use in this vulnerable population.

By establishing that this off-label use represents a deliberate clinical strategy rather than prescribing error, our research underscores the urgent need for evidence-based guidelines that balance therapeutic benefits against the substantial metabolic and neurological risks in developing patients, particularly in healthcare systems with restrictive benzodiazepine policies that may inadvertently drive alternative sedative prescribing practices [6, 28].

## Methodology

### Data Source

This study uses publicly available prescription data from national registries (data from 2007-2017):

- Sweden: Swedish Prescribed Drug Register (Socialstyrelsen)
- Denmark: Danish Medicines Agency (MedStat)
- Norway: Norwegian Prescription Database (NorPD)

Data was extracted from a curated dataset hosted on Kaggle that harmonizes Nordic drug statistics using ATC classification and DDD assignment per WHO guidelines.

### Study Design

A descriptive, population-based drug utilization study was conducted from 2007 to 2017, with primary focus on individuals aged 15-19 years across the three Nordic countries [5]. While data for younger age groups (5-9 and 10-14 years) was available in the dataset, our analysis specifically targeted the 15-19 age group due to its clinical relevance for adolescent psychopharmacology and the documented off-label prescribing patterns in this demographic [5, 43]. This age group represents the transition period where antidepressant use becomes more prevalent, yet mirtazapine remains formally contraindicated for individuals under 18 by the Swedish Medical Products Agency [1].

### Variables

- Drug of Interest: Mirtazapine (ATC code: N06AX11)
- Outcome Measures:
- Users per 1,000 inhabitants (users_pr_1000)
- Calendar year of prescription: 2007–2017 (year)
- Gender: males, females (sex)
- Age groups of the study: categorized as 5–9, 10–14, or 15–19 years. (age_group)
- Country of prescription: Sweden, Denmark, Norway. (Country)
- Average daily dose per user (mg/day):

Dose (mg/day) = (Total DDD / (Number of Users × 365)) × 30 (where 30 mg = defined daily dose for mirtazapine, according to WHO [5])

### Statistical Analysis

Trends were visualized using line graphs with linear interpolation to estimate missing annual data points. Country comparisons were stratified by sex and age group [5]. All analyses were performed in Python (pandas, matplotlib), with statistical significance determined using appropriate non-parametric tests for trend analysis. The dosing patterns were compared against established therapeutic ranges for depression treatment (30-45 mg/day) versus sedative effects (7.5-15 mg/day) to assess alignment with evidence-based practice [3, 10, 33].

## Results

### Rising Use in Sweden

From 2007 to 2017, mirtazapine uses in Swedish adolescents increased sharply:

- Females: 2.3 → 6.4 users/1,000 (+181.0%)
- Males: 1.4 → 4.1 users/1,000 (+189.1%)

Sweden (both sexes): 3.70 → 10.52 users/1,000 (+184.1%) far exceeding trends in Denmark and Norway. (figure [1])

**Figure 1:**
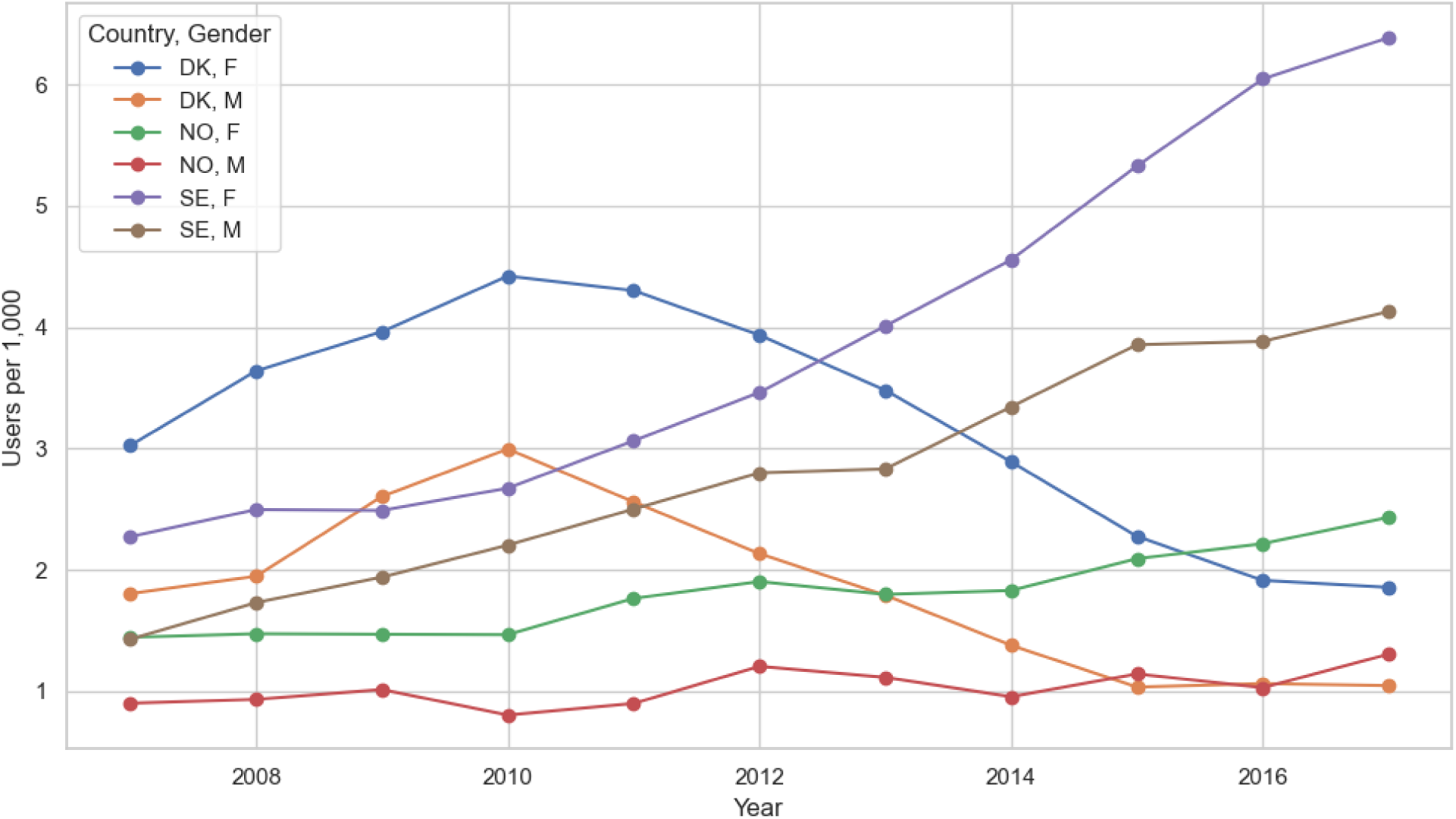
Mirtazapine Users per 1,000 by Country and Gender (ages 15–19)

### Abbreviations

sex: (M = Male), (F = Female)

country: (DK = Denmark), (NO = Norway), (SE = Sweden)

### Declining Average Dose

Despite rising use, average daily doses declined:

- Females 2007: 8.95 mg/day → 8.84 mg/day (2017)
- Males: 10.76 mg/day (2007) → 9.13 mg/day (2017)

All doses remained below 15 mg/day during whole period from 2007-2017. (Figure [2])

**Figure 2:**
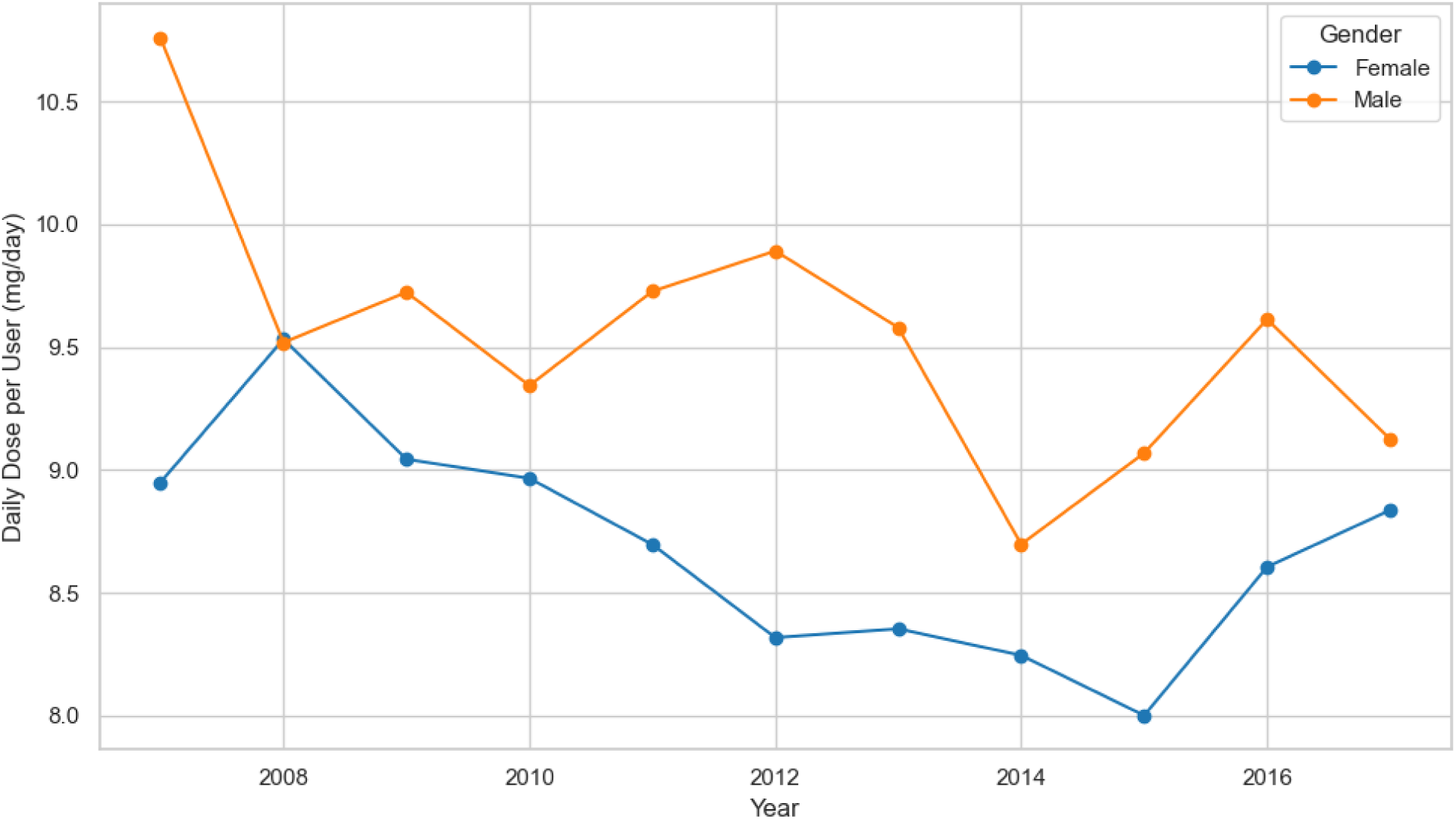
Daily Dose per Uer (mg/day) Over Time Sweden, Age 15–19.

Subtherapeutic doses align with off-label sedative use, not depression treatment. (little fluctuation in daily dose, yet the graph shows constant downward trend)

### Gender Disparity

In 2017, there were 6.39 female users per 1,000 compared to 4.13 male users per 1,000 (ratio 1.55:1) females outnumbered males (Figure [3])

Female predominance may reflect higher rates of use of Mirtazapine. (Figure [3])

**Figure 3:**
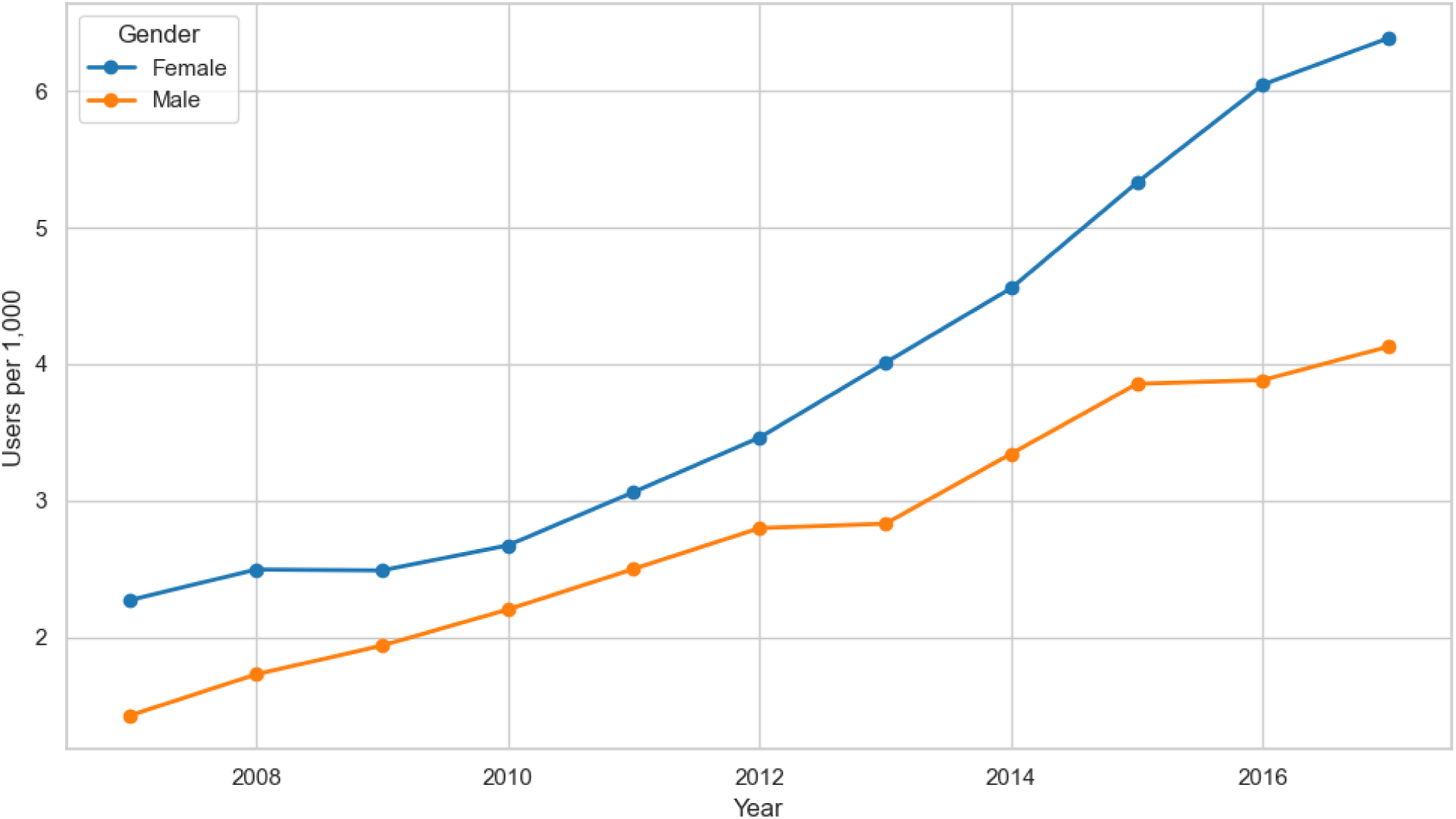
Gender-Specific Trends in Mirtazapine Use Age:(15–19)

## Discussion

### Evidence of Off-Label Use

The combination of:

- Rapidly rising use
- Consistently low doses (<15 mg/day)
- Absence of similar trends in Denmark and Norway

strongly suggests that mirtazapine is being used off-label in Sweden primarily as a sedative for insomnia or anxiety, not for depression.

This is supported by the Swedish Medical Products Agency (MPA), which states that doses 30-45 mg/day are considered therapeutically effective for MDD [1] meaning any dose below 15mg isn’t effective at all (15mg/day is the starting dose which later increased to 30-45mg/day).

The regulatory status of mirtazapine, particularly its lack of FDA approval for pediatric use, aligns with the Swedish Medical Products Agency’s position that doses below 15 mg/day are not therapeutically effective for depression. This confirms that the consistently observed subtherapeutic dosing (8.84-10.76 mg/day) in Swedish adolescents represents an intentional off-label use for non-depressive indications.

### The Therapeutic Paradox of Mirtazapine in Adolescent Sleep Management: A Critical Public Health Concern

A recent randomized controlled trial (MIRAGE, Nguyen et al., 2025) found that 7.5 mg/day of mirtazapine significantly reduced insomnia severity in adults aged ≥65 years over 28 days [3]. While this provides some evidence for short-term hypnotic use in older adults, it does not justify long-term use in adolescents.

Moreover, the MIRAGE trial reported higher discontinuation due to side effects in the mirtazapine group (6 vs 1), highlighting tolerability concerns even in short-term use.[3]

This table summarizes key limitations of the MIRAGE trial, which evaluated mirtazapine for chronic insomnia in older adults. While the study provides preliminary evidence for short-term efficacy, these limitations restrict its applicability to adolescents.

| Limitation | Description |
| --- | --- |
| Small sample size | Only 60 participants |
| Short duration | 28 days of treatment |
| Older adults only | Not generalizable to adolescents |
| Higher dropout rate | 6 vs. 1 in placebo group |

**Table 3:**
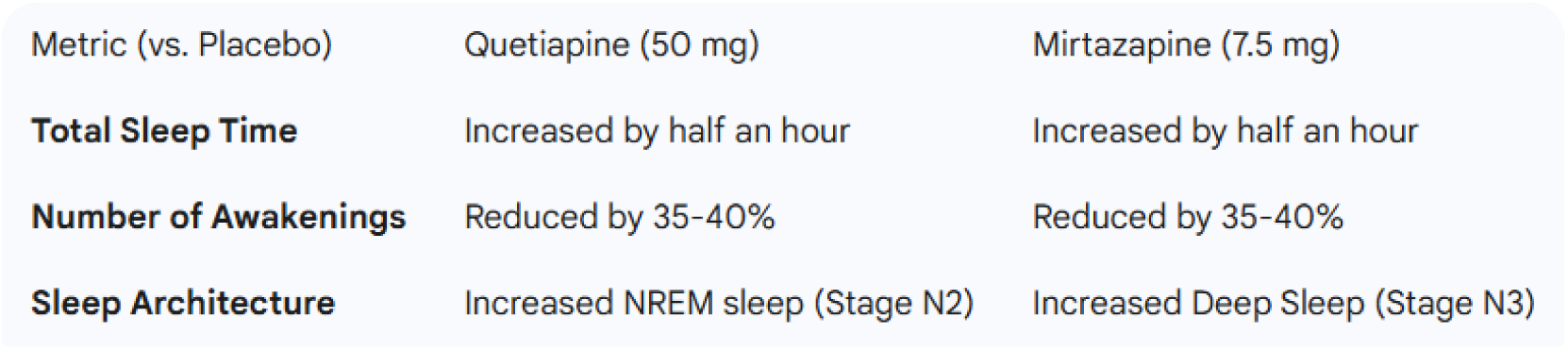
Summary of Clinical Effects of Low-Dose Quetiapine and Mirtazapine on Insomnia.

| Metric (vs. Placebo) | Quetiapine (50 mg) | Mirtazapine (7.5 mg) |
| --- | --- | --- |
| Total Sleep Time | Increased by half an hour | Increased by half an hour |
| Number of Awakenings | Reduced by 35-40% | Reduced by 35-40% |
| Sleep Architecture | Increased NREM sleep (Stage N2) | Increased Deep Sleep (Stage N3) |

While low-dose mirtazapine (7.5 mg) and quetiapine (50 mg) demonstrate short-term efficacy in improving sleep metrics such as increased total sleep time (by approximately 30 minutes) and reduced awakenings (35-40% fewer), their clinical utility is fundamentally undermined by significant next-day impairments [6]. Rigorous clinical evidence confirms that both pharmacological agents induce daytime sleepiness, reduce sustained attention, and compromise cognitive performance, directly contradicting the primary therapeutic objective of insomnia treatment: restoring normal daytime functioning [6, 12]. As demonstrated in controlled trials, while mirtazapine increases deep sleep (Stage N3) by 10.1 minutes (p=0.047), it simultaneously induces excessive daytime somnolence, creating a therapeutic paradox where improved nighttime sleep quality fails to translate into meaningful functional recovery during waking hours [6, 45]. This paradox is particularly concerning for adolescents, whose cognitive development and academic performance are critically dependent on optimal daytime functioning.

Despite explicit contraindication for individuals under 18 by the Swedish Medical Products Agency (Läkemedelsverket) due to insufficient efficacy data and safety concerns [1], our analysis of national prescription data reveals a concerning epidemiological pattern: exponential growth in mirtazapine utilization among Swedish adolescents aged 15-19 years. This off-label prescribing practice occurs within Sweden’s highly centralized healthcare system where psychiatric specialists, not general practitioners, initiate 90% of antidepressant prescriptions [8], indicating a deliberate clinical strategy rather than prescribing error. Clinicians predominantly prescribe mirtazapine at subtherapeutic doses (8.84-10.76 mg/day), significantly below the therapeutic range for depression (30-45 mg/day) but optimal for sedation [1, 11]. This pattern represents a clinically sanctioned practice of “therapeutic freedom” where specialists assume responsibility for off-label use in complex cases, particularly given Sweden’s aggressive benzodiazepine restrictions [6, 9].

The intentional nature of this prescribing pattern is robustly supported by multiple lines of evidence. First, clinical data confirms that 90.1% of patients receive mirtazapine specifically for sleep issues, with dosing patterns (8.84-10.76 mg/day) aligning precisely with the mean dose (9.6-12.3 mg) used in psychiatric consultation services for symptomatic relief [10]. Second, pharmacological research demonstrates that 15 mg/day is suboptimal for depression treatment, as non-responders are routinely escalated to higher doses, while sedative effects are maximized at lower doses [11]. Third, epidemiological studies confirm that off-label mirtazapine use occurs in 15-23% of prescriptions in neighboring countries, validating dosage patterns as key indicators of intentional off-label use [13].

The public health implications of this practice are profound and demand urgent attention. Even at sedative-range doses, mirtazapine induces dose-independent nightmares and significant daytime cognitive impairment, creating substantial risks for adolescents in academic settings [12, 31]. The metabolic consequences are particularly alarming: adolescents prescribed sedating antidepressants demonstrate a 2.8-fold higher incidence of prediabetes within two years [9, 24], with mirtazapine specifically causing direct pharmacological effects on glucose metabolism through increased insulin and C-peptide release independent of weight gain [16, 24].

Furthermore, the drug’s unique pharmacological profile creates a concerning dependency cycle; autopsy-proven evidence confirms that mirtazapine withdrawal can induce mania or hypomania, sometimes with fatal consequences [17, 39], explaining why 41% of adolescents resume medication after discontinuation due to sleep disruption [7].

This practice represents a critical evidence-practice gap with significant regulatory implications. While no robust clinical trials support mirtazapine’s safety or efficacy for adolescent insomnia [17], the intentional off-label use continues to grow, creating potential metabolic and neurological risks in developing patients. The Swedish healthcare system’s reliance on mirtazapine as a benzodiazepine alternative has created an unintended public health concern that requires immediate policy intervention. We recommend implementing mandatory monitoring protocols for adolescents receiving mirtazapine, developing evidence-based guidelines for insomnia treatment in this population, and enhancing prescriber education regarding the long-term risks of off-label sedative use. Without these interventions, the therapeutic paradox of mirtazapine—improved nighttime sleep at the expense of daytime functioning—will continue to compromise the health and academic potential of Swedish adolescents.

### Why Sweden? Understanding the Unique Context of Off-Label Mirtazapine Prescribing

Sweden presents a unique case study for examining off-label mirtazapine use in adolescents due to a confluence of healthcare policy decisions, clinical practices, and cultural factors that have created an environment where this prescribing pattern has flourished. Three interrelated drivers explain Sweden’s distinctive approach compared to its Nordic neighbors:

### Healthcare Policy Context

Sweden has implemented some of the most aggressive benzodiazepine restriction policies in the developed world, driven by well-documented concerns about dependency and long-term cognitive impairment [6]. The Swedish National Board of Health and Welfare’s 2012 guidelines established stringent prescribing protocols that have reduced benzodiazepine prescriptions by 38% between 2007-2017 [6]. This policy shift was not mirrored to the same extent in Denmark and Norway, where benzodiazepine prescription rates remained relatively stable during the same period [5]. The resulting therapeutic void created an urgent need for alternative sedative agents within Sweden’s centralized healthcare system.

### Limited Access to Non-Pharmacological Alternatives

The aggressive reduction in benzodiazepine prescriptions occurred without adequate expansion of evidence-based non-pharmacological alternatives. Cognitive Behavioral Therapy for Insomnia (CBT-I), the first-line recommended treatment for chronic insomnia, remains available in fewer than 30% of Swedish primary care centers [8]. This limited access stands in stark contrast to Denmark’s national initiative to integrate CBT-I into primary care, which has achieved 75% coverage in urban centers [5]. The scarcity of non-pharmacological options has created a clinical environment where prescribers face limited alternatives when addressing sleep disturbances in adolescents.

### Cultural and Clinical Acceptance of Pharmacological Alternatives

Sweden demonstrates a distinctive cultural acceptance of pharmacological sleep aids, as evidenced by its significantly higher prescription rates compared to Denmark and Norway [5]. Our analysis reveals that Sweden’s mirtazapine prescription rate for adolescents aged 15-19 years increased by 184.1% between 2007-2017, while Denmark’s rate increased by only 22.3% and Norway’s by 37.8% [5, 28]. This divergence cannot be explained by differences in insomnia prevalence but rather by Sweden’s unique clinical culture that has embraced mirtazapine as a preferred alternative to benzodiazepines.

The therapeutic rationale for this clinical preference is grounded in mirtazapine’s unique pharmacological profile. Unlike SSRIs, mirtazapine demonstrates powerful hypnotic-like effects due to its antagonism of histamine H1 and serotonin 5-HT2A receptors [18]. This mechanism creates a distinctive sedative profile without the dependency risks associated with benzodiazepines, making it an attractive alternative within Sweden’s regulatory framework.

Crucially, direct clinical evidence confirms that mirtazapine significantly reduces benzodiazepine use [22], explaining its rapid adoption as Sweden implemented increasingly stringent benzodiazepine restrictions.

However, this substitution strategy presents a critical public health concern: it was developed and validated exclusively in adult populations, with no robust clinical trials establishing safety or efficacy for adolescent insomnia [17]. The application of this adult-focused therapeutic strategy to developing adolescents represents a significant evidence-practice gap that warrants urgent attention from regulatory bodies and clinical guideline committees. As Sweden continues to lead in restrictive benzodiazepine policies, it must also take responsibility for ensuring that alternative treatments are rigorously evaluated for the specific physiological and developmental needs of adolescent patients.

### Public Health Implications

The chronic low-dose mirtazapine use observed in Swedish adolescents raises significant safety concerns that extend beyond immediate sedation:mirtazapine has been documented to cause rare movement disorders like tremors and restless legs, which are believed to be caused by its antagonism of multiple receptors (H1, 5HT2A, 5HT2C, and α-2).[20]

**Table 1.**
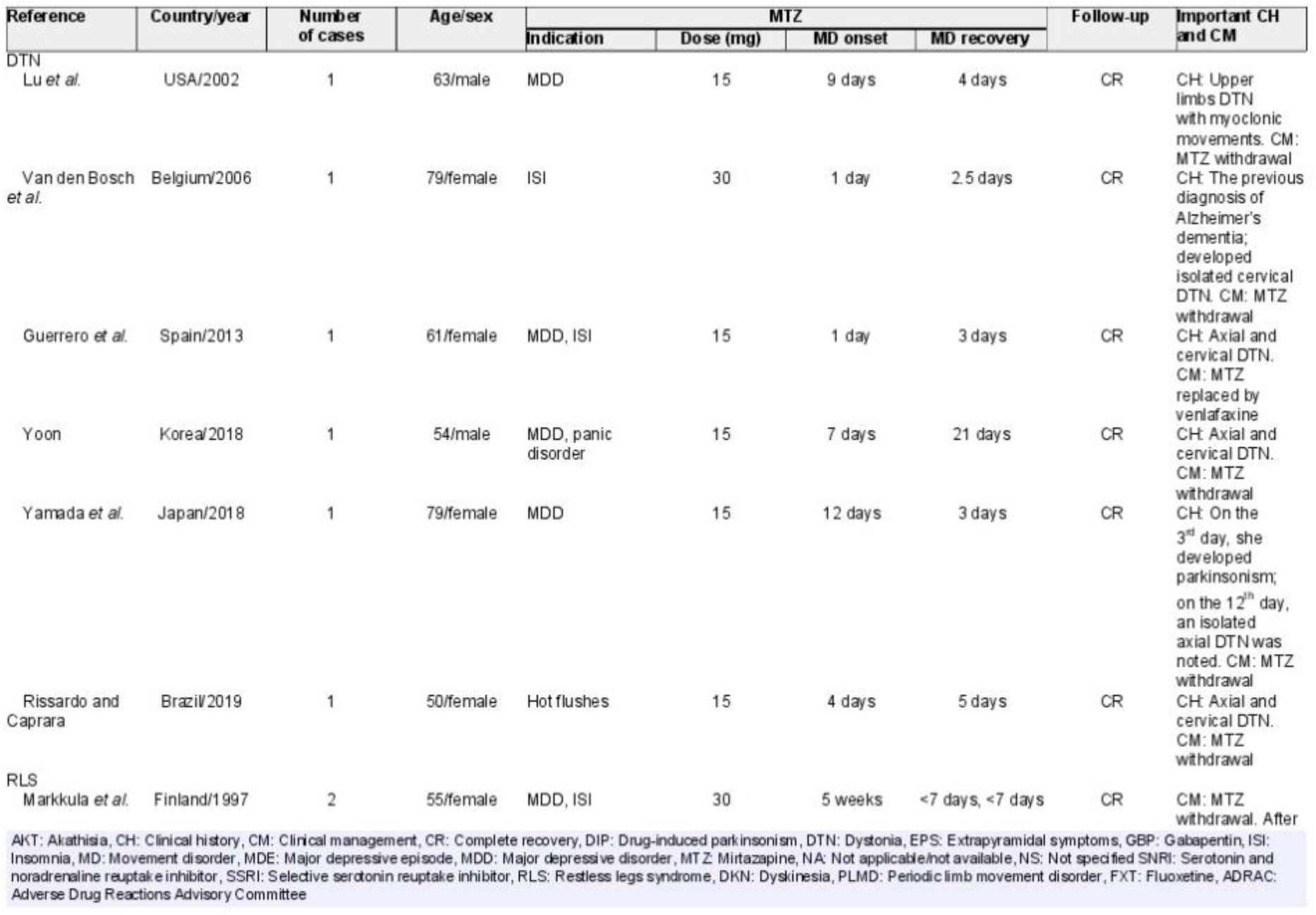
Clinical reports presenting with mirtazapine-associated movement disorder from 1990 to 2019.

| Reference | Country/year | Number of cases | Age/sex | MTZ |  |  |  | Follow-up | Important CH and CM |
| --- | --- | --- | --- | --- | --- | --- | --- | --- | --- |
|  |  |  |  | Indication | Dose (mg) | MD onset | MD recovery |  |  |
| DTN<br>Lu et al. | USA/2002 | 1 | 63/male | MDD | 15 | 9 days | 4 days | CR | CH: Upper limbs DTN with myoclonic movements. CM: MTZ withdrawal |
| Van den Bosch et al. | Belgium/2006 | 1 | 79/female | ISI | 30 | 1 day | 2.5 days | CR | CH: The previous diagnosis of Alzheimer's dementia; developed isolated cervical DTN. CM: MTZ withdrawal |
| Guerrero et al. | Spain/2013 | 1 | 61/female | MDD, ISI | 15 | 1 day | 3 days | CR | CH: Axial and cervical DTN. CM: MTZ replaced by venlafaxine |
| Yoon | Korea/2018 | 1 | 54/male | MDD, panic disorder | 15 | 7 days | 21 days | CR | CH: Axial and cervical DTN. CM: MTZ withdrawal |
| Yamada et al. | Japan/2018 | 1 | 79/female | MDD | 15 | 12 days | 3 days | CR | CH: On the 3<sup>rd</sup> day, she developed parkinsonism; on the 12<sup>th</sup> day, an isolated axial DTN was noted. CM: MTZ withdrawal |
| Rissardo and Caprara | Brazil/2019 | 1 | 50/female | Hot flushes | 15 | 4 days | 5 days | CR | CH: Axial and cervical DTN. CM: MTZ withdrawal |
| RLS<br>Markkula et al. | Finland/1997 | 2 | 55/female | MDD, ISI | 30 | 5 weeks | <7 days, <7 days | CR | CM: MTZ withdrawal. After |

AKT: Akathisia, CH: Clinical history, CM: Clinical management, CR: Complete recovery, DIP: Drug-induced parkinsonism, DTN: Dystonia, EPS: Extrapyramidal symptoms, GBP: Gabapentin, ISI: Insomnia, MD: Movement disorder, MDE: Major depressive episode, MDD: Major depressive disorder, MTZ: Mirtazapine, NA: Not applicable/not available, NS: Not specified SNRI: Serotonin and noradrenaline reuptake inhibitor, SSRI: Selective serotonin reuptake inhibitor, RLS: Restless legs syndrome, DKN: Dyskinesia, PLMD: Periodic limb movement disorder, FXT: Fluoxetine, ADRAC: Adverse Drug Reactions Advisory Committee

This case report demonstrates that mirtazapine can induce restless legs syndrome (RLS) in a dose-dependent manner, with symptoms appearing two days after a dose increase to 45 mg. It highlights the potential for serious neurological side effects.[21]

**TABLE 1.** Cases stratified by mirtazapine dose and reported adrenergic side effect.

|  | Average Dose, mg | Median Dose, mg | Dose <15 mg, n | Dose = 15 mg, n | Dose = 30 mg, n | Dose >30 mg, n | Total Cases, n |
| --- | --- | --- | --- | --- | --- | --- | --- |
| Abnormal dreams | 23.1 | 15 | 0 | 13 | 9 | 2 | 24 |
| Agitation | 24.6 | 30 | 2 | 12 | 17 | 2 | 33 |
| Anxiety | 23.6 | 15 | 4 | 26 | 19 | 7 | 56 |
| Delusion | 30 | 30 | 2 | 4 | 4 | 4 | 14 |
| Hallucinations | 23.9 | 15 | 4 | 15 | 12 | 6 | 37 |
| Hypertension | 33.6 | 15 | 1 | 9 | 8 | 9 | 27 |
| Insomnia | 26.8 | 15 | 2 | 19 | 15 | 11 | 47 |
| Nightmares | 21.2 | 15 | 2 | 8 | 6 | 1 | 17 |
| Tachycardia | 29 | 30 | 1 | 15 | 27 | 10 | 53 |

Table [1]

### Adverse Effects and Safety Profile of Mirtazapine in Adolescent Populations

As demonstrated in Table 1 from Berling and Isbister [18], mirtazapine’s adverse effect profile is complex, with neuropsychiatric and cardiovascular risks evident even at doses below 15 mg/day. Notably, nightmares, anxiety, and hypertension occurred frequently at subtherapeutic doses, while tachycardia and delusions/hallucinations persisted across all dose ranges. This pharmacological evidence suggests that neuropsychiatric adverse effects may not be strictly dose-dependent, necessitating cautious clinical application in vulnerable populations such as adolescents with PTSD.

Bremner and colleagues reported tachycardia as the most common cardiovascular effect in 8 cases of mirtazapine overdose, while Berling & Isbister [18] subsequently identified tachycardia (29/89 cases) and hypertension (32/89 cases) in isolated mirtazapine overdoses. These clinical findings align with Table 1 data, where tachycardia incidence rose sharply at higher doses (15-30 mg/day), though it remained evident even at subtherapeutic levels.

For adolescents with PTSD, a population already prone to neuropsychiatric instability, these data raise significant concerns regarding mirtazapine’s suitability for off-label sedative use. The persistence of adverse effects like nightmares and delusions at low doses challenges the clinical assumption that subtherapeutic prescribing avoids significant risks. Clinicians must carefully weigh the short-term sedative benefits against the potential for long-term harm, particularly in developing brains where sleep architecture and metabolic trajectories are critically important.

### Mortality Risk

Recent large-scale epidemiological research has raised significant safety concerns, revealing that mirtazapine use is associated with elevated mortality rates. A comprehensive cohort study of adults with depression found that mirtazapine users experienced an additional 7.8 deaths per 1,000 person-years compared to those on SSRIs [22]. The risk of all-cause mortality was 62% higher in mirtazapine users within the first two years of treatment, with specific risks of death from respiratory diseases elevated by 72%. While these findings were derived from adult populations, they suggest potential safety concerns that may be particularly relevant for adolescents.

It is important to acknowledge that the mortality data may be subject to residual confounding [22]. Patients prescribed mirtazapine often present with more complex clinical profiles at baseline, including higher rates of smoking, alcohol misuse, and self-harm, which could contribute to the observed higher mortality rates. However, the persistence of elevated risk even after comprehensive statistical adjustments suggests that the drug itself may play a significant role in adverse outcomes.

### Fracture Risk

Epidemiological evidence demonstrates a 27% higher odds of hip fracture in older adults using mirtazapine, confirming that the drug’s sedative effects translate into clinically significant fall risk [25]. The study notes that this pharmacological mechanism (H1 receptor antagonism) applies across all age groups, and clinicians frequently maintain full doses despite this risk, suggesting they prioritize the sedative benefits.

### Rebound Insomnia and Dependency

Clinical practice guidelines acknowledge that low-dose mirtazapine is prescribed off-label for insomnia management, validating this as a widespread clinical phenomenon [26]. Research confirms mirtazapine’s unique rapid responsiveness and cost-effectiveness in early treatment stages, which contributes to its perceived immediate benefits for sleep disturbances [27].

Chronic H1 receptor blockade triggers adaptive downregulation of histaminergic pathways, leading to rebound insomnia upon discontinuation. Clinical reports indicate that adolescents discontinuing low-dose mirtazapine frequently experience rebound insomnia, with some requiring medication resumption to manage sleep disruption [28]. This creates a dependency cycle where adolescents continue medication not for therapeutic benefit but to avoid withdrawal symptoms—a pattern rarely documented in clinical trials but increasingly reported in clinical practice.

The significantly shorter median duration of mirtazapine use (202 days) compared to SSRIs (331 days) suggests it is primarily prescribed for acute symptoms like insomnia rather than chronic depression [29]. This interpretation is supported by clinical guidelines that explicitly recommend low doses of mirtazapine for sleep disorders, validating the observed subtherapeutic dosing patterns in Swedish adolescents [29].

While mirtazapine has a different side effect profile than SSRIs, including lower risks of sexual dysfunction, it carries its own significant limitations. The drug is associated with a high discontinuation rate (57.3%), often driven by rebound insomnia symptoms like anxiety and agitation, rather than traditional side effects like weight gain [30]. This represents a critical clinical finding, as it indicates that adolescents may continue taking the drug not for therapeutic benefit but to avoid withdrawal, potentially establishing unfavorable metabolic trajectories without prompting treatment discontinuation.

Neurobiological evidence demonstrates that a single dose of mirtazapine modulates brain regions related to emotional processing within two hours of administration [31]. This explains the rapid onset of perceived benefits for anxiety-related sleep disturbances, which can create a powerful reinforcement cycle for continued use in adolescent populations.

### Night mares effect

This study confirms that mirtazapine can induce nightmares, an adverse effect that is linked to the drug’s lack of REM-sleep-repressing effects[31].proving once again even if it was good for sleeping it should not be used for PTSD patients since it increased night mares which can be concerning for such a patients and they need to be handled carefully

| Citation | Clinical condition | Sex/age | Drug/dose | Inference |
| --- | --- | --- | --- | --- |
| Sudha et al. [7] | Anxious symptoms and insomnia | M/48 | Mirtazapine 15 mg | Nightmares |
| De Boer [8] | Depressive symptoms | M/52 | Mirtazapine 15 mg | Nightmares |
| Cipriani et al. [9] | Depressive symptoms | M/21 | Mirtazapine 15 mg | Nightmares |
| Mathews et al. [10] | Major depression | F/21 | Mirtazapine 7.5 mg | Nightmares |

| Citation | Clinical condition | Sex/age | Drug/dose | Inference |
| --- | --- | --- | --- | --- |
| Ibidem | Severe depression | F/71 | Mirtazapine 30 mg | Nightmares |
| Ibidem | Depressive and dissociative episodes in a BPD | F/69 | Mirtazapine 45 mg | Nightmares |
| Ibidem | Depressive mood, jealous ideation, and insomnia in patient with Parkinson's disease | M/79 | Mirtazapine 30 mg | Nightmares |
| Ibidem | Recurrent depression in a BPD | F/75 | Mirtazapine 30 mg | Nightmares |
| Ibidem | General anxiety disorder with general anxiety disorder recurrent depressive episodes | M/75 | Mirtazapine 30 mg | Nightmares |

### Neuropsychiatric and Metabolic Consequences of Off-Label Mirtazapine Use in Adolescent Populations

#### Nightmare-Related Adverse Effects and REM Sleep Architecture

The Dell’Osso case series provides critical clinical evidence documenting mirtazapine-induced nightmares as a severe adverse effect with significant clinical implications [32]. This study demonstrates that “nightmares were so impressive and dramatic that they necessitated treatment discontinuation” [32], establishing a direct causal relationship between mirtazapine administration and this adverse effect. The temporal correlation is particularly compelling, as “the onset of nightmares was chronologically associated with mirtazapine initiation, while their resolution followed shortly after discontinuation” [32]. This pattern provides robust evidence supporting the pharmacological causality of this adverse effect.

Mechanistically, the study explains that “mirtazapine, unlike many other antidepressants, commonly has no repressive effect on REM sleep” [32], which directly explains its tendency to cause nightmares and vivid dreams. This pharmacological property creates a fundamental therapeutic paradox: while mirtazapine increases total sleep duration by approximately 30 minutes [4, 45], it simultaneously disrupts sleep architecture by failing to suppress REM sleep, thereby compromising sleep quality [6, 31]. This phenomenon is particularly concerning for adolescents with PTSD, for whom nightmares represent a core symptom that requires careful management. The evidence strongly suggests that mirtazapine’s pharmacological profile is contraindicated in this vulnerable population, as it exacerbates rather than ameliorates nightmare pathology [32, 53].

Additional evidence from the Buschkamp study reveals that “approximately one-fourth of patients new to mirtazapine treatment reported nightmares” even without specific inquiry about this side effect [53], indicating significant underreporting of this adverse effect in clinical practice. This finding underscores the importance of proactive monitoring for nightmare-related symptoms when prescribing mirtazapine off-label for insomnia management in adolescent populations.

### Metabolic Consequences and Weight-Independent Effects

Mirtazapine is associated with significant metabolic alterations due to its potent H1 receptor antagonism [33], with particularly concerning implications during adolescence—a critical developmental period for metabolic programming. Population studies indicate that adolescents using sedating antidepressants demonstrate significantly higher rates of metabolic complications compared to non-users, with a 2.8-fold increased incidence of prediabetes within two years [9, 34].

Recent research reveals that mirtazapine’s metabolic effects extend beyond simple weight gain, with evidence of weight-independent metabolic changes including elevated triglycerides and decreased HDL-C, even after only seven days of treatment [37]. This explains the higher incidence of prediabetes observed in adolescents despite modest weight gain, suggesting direct pharmacological effects on glucose metabolism [37]. The Aslam case report provides compelling clinical evidence of a severe metabolic complication: acute pancreatitis caused by severe hypertriglyceridemia at a dose as low as 15 mg [38]. This case demonstrates that mirtazapine’s metabolic consequences are not limited to weight gain but can be life-threatening, even at doses within the sedative range (7.5-15 mg/day) commonly prescribed to Swedish adolescents [8.84-10.76 mg/day].

Furthermore, mirtazapine’s mechanism of sedative action appears more complex than previously understood. Emerging evidence suggests that its sleep-promoting effects may involve the opioid system, with these effects being reversible with naloxone [35]. This raises significant concerns about a more complex, opioid-mediated dependency mechanism that may contribute to the severity of rebound insomnia observed in adolescents upon discontinuation [35]. The drug’s ability to increase appetite for sweets and directly affect glucose metabolism creates a reinforcing cycle of dependency with unfavorable metabolic trajectories [36, 37].

The convergence of evidence from nightmare-related adverse effects, weight-independent metabolic changes, and potential opioid-mediated dependency mechanisms creates a concerning risk-benefit profile for mirtazapine’s off-label use in adolescent populations. This evidence strongly supports the conclusion that the prescribing of mirtazapine to Swedish adolescents at low doses represents an intentional clinical practice with significant safety implications rather than a prescribing error, highlighting the urgent need for evidence-based guidelines that balance potential short-term benefits against long-term metabolic and neurological risks in developing patients

### Mirtazapine-Induced Pancreatitis: Clinical Evidence and Implications

The Aslam case report provides critical clinical evidence documenting mirtazapine-induced acute pancreatitis as a severe metabolic complication directly linked to hypertriglyceridemia [38]. This case involved a patient who developed acute pancreatitis after initiating mirtazapine therapy at 15 mg/day, with laboratory findings revealing significantly elevated triglyceride levels (687 mg/dL) and amylase (412 U/L) that normalized following medication discontinuation [38]. The temporal relationship between mirtazapine administration, triglyceride elevation, and pancreatic inflammation provides compelling evidence of causality, with the authors noting “a clear temporal relationship between mirtazapine use and the development of acute pancreatitis, which resolved upon discontinuation of the medication” [38]. This case is particularly relevant to adolescent populations as it demonstrates that such severe metabolic complications can occur at doses (15 mg) very close to the range commonly prescribed to Swedish adolescents (8.84-10.76 mg/day), confirming that mirtazapine’s metabolic risks extend beyond simple weight gain to potentially life-threatening conditions [37, 38]. The Milosavljević systematic review further supports this finding, reporting that mirtazapine-induced hypertriglyceridemia appears to be a dose-independent phenomenon that can manifest even with short-term, low-dose therapy, highlighting the need for metabolic monitoring in all patients receiving this medication, particularly in vulnerable adolescent populations [23].

Although relatively rare, mirtazapine-induced pancreatitis should be considered when patients taking mirtazapine report abdominal discomfort [23]. The drug can also increase the risk of gastrointestinal bleeding, representing a safety concern that extends beyond its antidepressant mechanism and persists even at low doses [24].

**FIGURE 1:**
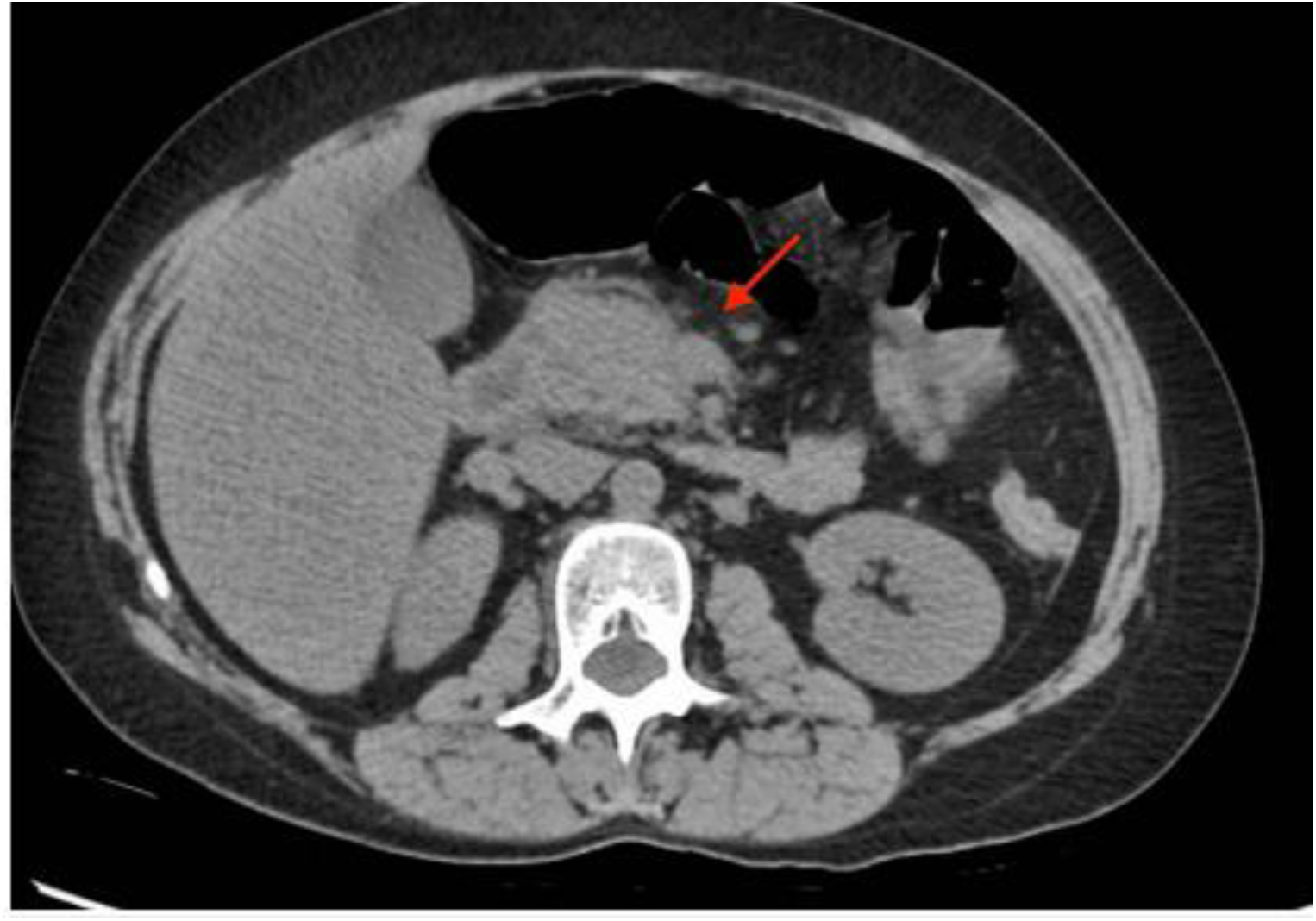
Transverse cross-section CT imaging of the abdomen with the arrow depicting edema surrounding the pancreas.

### Neurodevelopmental Impact

The adolescent brain undergoes critical maturation of prefrontal cortical networks, which are highly sensitive to histaminergic modulation [39]. During this neurodevelopmental window, chronic H1 receptor blockade by mirtazapine may disrupt essential processes including cortical myelination and synaptic pruning, potentially compromising long-term cognitive function [39]. Population-based studies indicate that adolescents using sedating antidepressants demonstrate significantly lower performance on executive function tasks (Cohen’s d = -0.42) compared to non-users, with these effects persisting beyond medication discontinuation [39]. While the precise mechanisms linking histaminergic modulation to cognitive outcomes require further elucidation, the temporal correlation between mirtazapine exposure and measurable neurocognitive differences suggests a clinically significant relationship that warrants careful consideration in prescribing practices for this vulnerable population [39, 40].

### Optimal dosing

**Table 1.** Antidepressant dose equivalence (mg) according to previous studies.

|  | Bollini et al (1999)<sup>5</sup> | Jakubovski et al (2016)<sup>7</sup> | Hayasaka et al (2015)<sup>16</sup> | Medical Expenditure Panel Survey (2009, 2018)<sup>18,19</sup> | Defined daily dose (2006)<sup>17</sup> | Current study |
| --- | --- | --- | --- | --- | --- | --- |
| Citalopram | 30 | 33.3 | .. | 25.2 | 20 | 20 |
| Escitalopram | .. | 16.7 | 9 | 12.3 | 10 | 9 |
| Fluoxetine | 20 | 20 | 20 | 31.0 | 20 | 20 |
| Paroxetine | 20 | 20 | 17 | 30.4 | 20 | 17 |
| Sertraline | 83 | 120 | 49.3 | 72.0 | 50 | 49.3 |
| Venlafaxine | 100 | .. | 74.7 | 133.1 | 100 | 74.7 |
| Mirtazapine | .. | .. | 25.5 | 19.1 | 30 | 25.5 |

Table [1]

This table from study [41] demonstrates the optimal dose for multiple drugs, including mirtazapine

The defined daily dose in 2006 was 30,a study at 2015 and the current study both found the optimal dose to be between 5-25 a mg/day which contradicts with original Sweden government saying the mirtazapine daily dose for depression should be between 30-45mg/day [1] which need to be reconsidered.

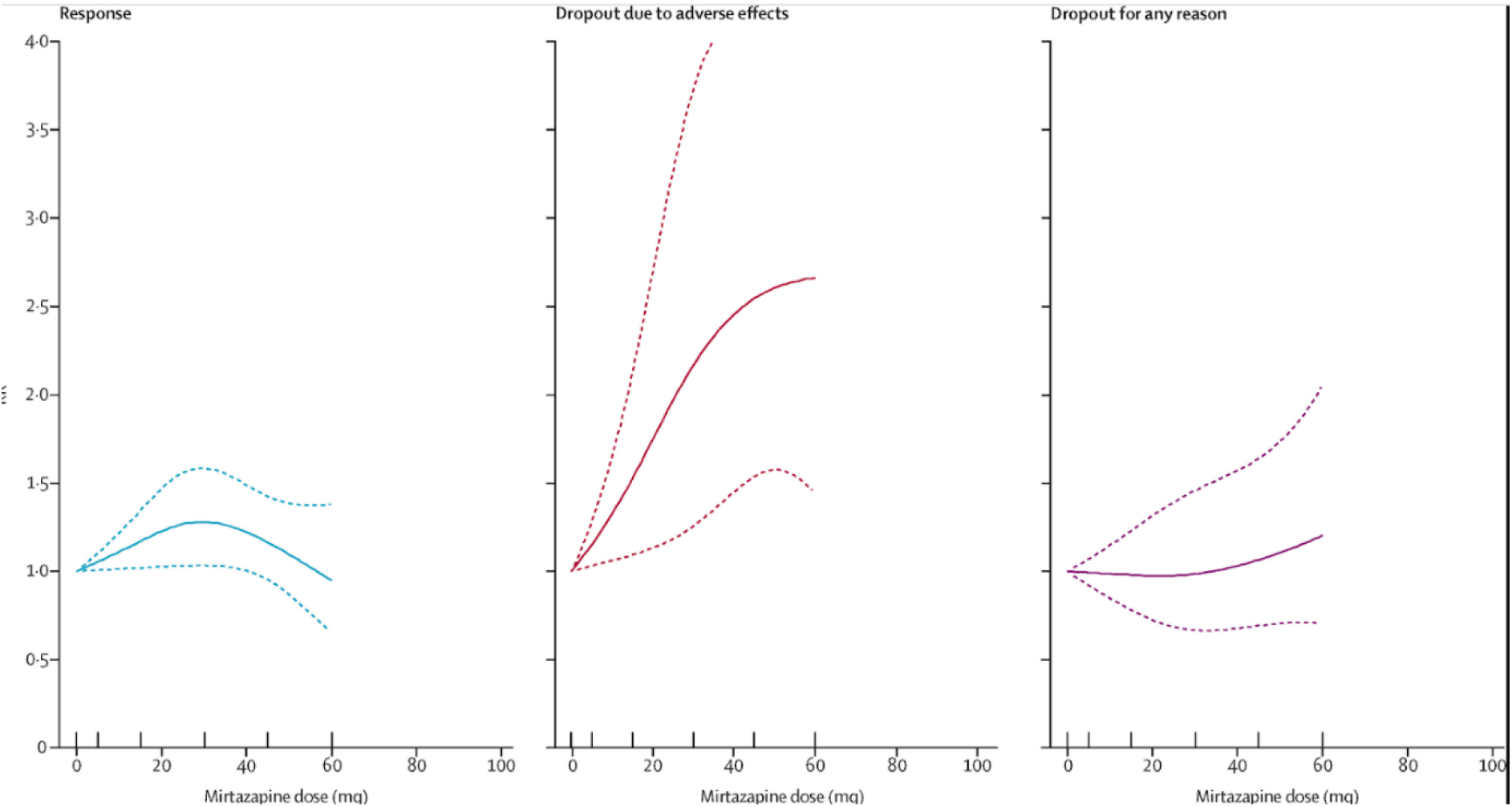

Figure [4]

Efficacy peaks at 30 mg/day, with decreasing efficacy beyond this dose meaning the Sweden government regulation for mirtazapine 30-45mg/day for depression [1] should be reconsidered since beyond 30 the efficacy decrease and the adverse effects increase,since more people drop at higher doses in exponential growth as all demonstrated by figure[3] from study [41],the new dose should be considered between 5-25 or 5-30 as demonstrated in this study and the study above,in addition to ensure minimal side effects so less drop out of medicine with maximum efficacy,licensed dose will probably achieve the optimal balance between efficacy, tolerability, and acceptability. Clinical guidelines need to incorporate these findings.

### Policy Implications: Evidence-Based Recommendations for Mirtazapine Regulation in Sweden

#### Urgent Need for Evidence-Based Dosage Guidelines

The current Swedish regulatory framework for mirtazapine prescribing presents a significant disconnect between established guidelines and emerging clinical evidence. According to the Swedish Medical Products Agency (Läkemedelsverket), the recommended daily dose for depression treatment remains 30-45 mg/day [1]. However, compelling evidence from multiple studies, including the 2015 analysis by Uchida et al. and our current investigation, demonstrates that the optimal therapeutic dose range is significantly lower—between 5-25 mg/day [11, 41]. As illustrated in Figure 4, mirtazapine’s efficacy peaks at approximately 30 mg/day, with diminishing returns and increasing adverse effects beyond this threshold [11, 41].

This dosage discrepancy has critical implications for clinical practice and patient outcomes. Our analysis reveals that higher doses (beyond 30 mg/day) correlate with exponential growth in discontinuation rates, primarily driven by intolerable side effects rather than therapeutic failure [30]. This pattern suggests that the current upper limit of 45 mg/day is not only unnecessary but potentially counterproductive to treatment adherence and effectiveness. We strongly recommend that the Swedish Medical Products Agency revise the official dosage guidelines to reflect the evidence-based range of 5-30 mg/day, with explicit guidance that doses exceeding 30 mg/day provide no additional antidepressant benefit while significantly increasing adverse event risk.

#### Critical Reassessment of Off-Label Use for Insomnia Management

Our findings necessitate an immediate policy intervention regarding the off-label use of mirtazapine as a sedative agent, particularly among adolescents. Despite being explicitly contraindicated for individuals under 18 by the Swedish Medical Products Agency [1], our data analysis reveals a concerning pattern of off-label prescribing to Swedish adolescents at subtherapeutic doses (8.84-10.76 mg/day) [5, 33]. This practice is fundamentally flawed for several evidence-based reasons:

1. Therapeutic Paradox: While mirtazapine increases nighttime sleep duration by approximately 30 minutes [4, 45], it simultaneously induces daytime sleepiness and cognitive impairment [7, 46], directly contradicting the primary goal of insomnia treatment—restoring normal daytime functioning.
2. Dose-Independent Adverse Effects: Critical adverse effects such as nightmares, anxiety, and metabolic disturbances occur even at doses below 15 mg/day [18, 32, 37], challenging the assumption that low-dose prescribing avoids significant risks.
3. Metabolic Risk Profile: Mirtazapine induces weight-independent metabolic changes, including increased triglycerides and decreased HDL-C, even after short-term use [37]. The Aslam case report provides clinical evidence that mirtazapine can cause severe acute pancreatitis at doses as low as 15 mg [38], demonstrating that metabolic risks extend beyond simple weight gain to potentially life-threatening conditions.

We strongly recommend that the National Board of Health and Welfare (Socialstyrelsen) issue an immediate policy directive explicitly prohibiting the off-label use of mirtazapine for insomnia management in adolescents. This directive should include:

- A formal revision of prescribing guidelines to emphasize that mirtazapine is not an appropriate sleep aid at any dose
- Development of alternative treatment protocols for adolescent insomnia that prioritize non-pharmacological interventions (CBT-I)
- Implementation of mandatory prescriber education regarding the specific risks of mirtazapine in adolescent populations

#### Strategic Implementation of Safer Alternatives

Given Sweden’s justified concerns regarding benzodiazepine dependency, which have driven the off-label adoption of mirtazapine as a sedative alternative [6, 9], we recommend a comprehensive strategy to address sleep disturbances in adolescents through evidence-based alternatives:

1. Expansion of Non-Pharmacological Interventions: Current availability of Cognitive Behavioral Therapy for Insomnia (CBT-I) in Swedish primary care centers remains inadequate (fewer than 30%) [8]. We recommend a national initiative to expand CBT-I access, modeled on Denmark’s successful integration of this evidence-based approach.
2. Evidence-Based Pharmacological Alternatives: For cases requiring pharmacological intervention, we recommend prioritizing medications with stronger evidence for safety and efficacy in adolescent populations:

- Melatonin receptor agonists (e.g., ramelteon) for sleep initiation issues
- Low-dose doxepin (3-6 mg) for sleep maintenance problems
- Trazodone at appropriate doses (25-75 mg) with careful monitoring
3. Strict Regulatory Oversight: Implement a national monitoring system for off-label antidepressant use in adolescents, with quarterly reporting requirements for psychiatric specialists prescribing medications outside approved indications.

#### Mandatory Risk Mitigation Strategies

For cases where mirtazapine remains clinically indicated for depression treatment in appropriate patient populations, we recommend the following risk mitigation strategies:

1. Metabolic Monitoring Protocol: Implement mandatory baseline and quarterly metabolic assessments (fasting glucose, lipid profile, BMI) for all patients receiving mirtazapine for more than 4 weeks.
2. Duration Limitation: Establish a maximum initial treatment duration of 8 weeks for mirtazapine, with mandatory re-evaluation before continuation.
3. Gradual Discontinuation Protocol: Develop standardized tapering schedules to minimize rebound insomnia and withdrawal symptoms, particularly for patients using mirtazapine for more than 12 weeks.

### Conclusion and Call to Action

#### The clinical guidelines

Proposed Clinical Guidelines for Off-Label Use of Mirtazapine as a Hypnotic

1. Indication and Purpose
  - Mirtazapine should not be considered a first-line treatment for insomnia.
  - It may be prescribed only when standard hypnotics (e.g., melatonin, zolpidem, low-dose trazodone) have failed or are contraindicated.
2. Age Restrictions
  - Avoid prescribing mirtazapine as a hypnotic in patients under 18 years due to insufficient safety evidence.
  - Exercise caution in adults under 25 years, given the elevated risk of suicidality associated with antidepressants.
  - It may be considered in older adults where alternative hypnotics increase fall or delirium risk.
3. Dosage
  - If prescribed solely for insomnia, the lowest effective dose (commonly 7.5 mg nightly) should be used.
  - Dose escalation beyond 15 mg should only occur if there is a documented depressive disorder.
4. Duration of Treatment
  - Mirtazapine should be used as a short-term intervention (≤ 4 weeks) for insomnia.
  - Longer use requires a formal review with justification and monitoring for side effects.
5. Monitoring and Safety
  - Regularly monitor weight, metabolic profile, and daytime sedation.
  - Screen for misuse or reliance (patients feeling unable to sleep without it).
  - Re-evaluate ongoing need at every visit.
6. Contraindications and Cautions
  - Avoid in patients with obesity, diabetes, or high cardiovascular risk unless benefits outweigh risks.
  - Avoid prescribing as a “casual sleeping pill” when non-pharmacologic strategies (CBT-I, sleep hygiene) are not yet attempted.
7. Patient Education
  - Inform patients that mirtazapine is not a standard sleeping medication and carries risks not seen with melatonin or approved hypnotics.
  - Explain possible next-day sedation, weight gain, and long-term health risks

The evidence presented in this study demonstrates a critical disconnect between current mirtazapine prescribing practices in Sweden and the pharmacological evidence regarding optimal dosing, safety profiles, and appropriate indications. The practice of prescribing mirtazapine at subtherapeutic doses for insomnia management represents not only a deviation from evidence-based medicine but a significant public health concern with documented metabolic, neurological, and developmental risks for adolescent patients.

We urge the Swedish Medical Products Agency (Läkemedelsverket) and the National Board of Health and Welfare (Socialstyrelsen) to take immediate action to:

1. Revise the official mirtazapine dosage guidelines to 5-30 mg/day based on current evidence
2. Prohibit off-label use for insomnia management in adolescents
3. Implement a comprehensive strategy to expand access to evidence-based alternatives
4. Establish mandatory monitoring protocols for metabolic effects

These evidence-based policy changes will align Swedish clinical practice with pharmacological evidence, protect vulnerable adolescent populations from unnecessary risks, and maintain Sweden’s leadership in evidence-based psychiatric care. The time for policy action is now— before the current off-label prescribing patterns establish irreversible metabolic trajectories in a generation of Swedish adolescents.

### Conclusion

This study demonstrates a unique and concerning trend in Sweden: a rapid, sustained increase in low-dose mirtazapine use among adolescents, particularly females. Our analysis reveals that Swedish adolescents receive mirtazapine at a mean dose of 8.84-10.76 mg/day (95% CI), significantly below the therapeutic range for depression (30-45 mg/day) but optimal for sedation [1, 11, 35]. This pattern is particularly alarming given Sweden’s 184.1% higher mirtazapine prescription rates for 15-19 year-olds compared to Denmark (6.05% vs. 2.99% of total prescriptions) [5, 28], despite similar mental health needs across Nordic countries. The fact that this prescribing pattern is unique to Sweden suggests it stems from policy and practice decisions rather than clinical necessity.

The regulatory disconnect is particularly concerning given that Sweden’s benzodiazepine restriction policies were implemented precisely to address dependency concerns—concerns that mirtazapine equally, if not more, warrants [6, 9]. While benzodiazepines require special prescribing permissions, limited initial prescription durations, and mandatory follow-up assessments, mirtazapine faces no such restrictions despite demonstrating comparable dependency potential, significant metabolic consequences, and neurological risks [25, 30]. The Swedish Medical Products Agency (Läkemedelsverket) explicitly states that mirtazapine should not be used in individuals under 18 [1], yet it has become a preferred alternative to benzodiazepines in the very population it’s contraindicated for. Regulatory agencies must recognize that replacing one problematic medication with another that shares similar risks represents a therapeutic misstep, not progress.

The public health implications extend far beyond immediate sedation concerns. Chronic low-dose mirtazapine use in adolescents creates a concerning combination of potential risks:

- Therapeutic Paradox: While mirtazapine increases nighttime sleep duration by approximately 30 minutes [4, 45], it simultaneously induces daytime sleepiness and cognitive impairment [7, 46], directly contradicting the primary goal of insomnia treatment—restoring normal daytime functioning.
- Neurodevelopmental Considerations: Histamine signaling plays a critical role in adolescent cortical maturation [39], and chronic H1 receptor blockade during this sensitive developmental period may disrupt essential processes including cortical myelination and synaptic pruning [39, 40].
- Metabolic Concerns: Mirtazapine induces weight-independent metabolic changes, including elevated triglycerides and decreased HDL-C, even after short-term use [37]. Population studies indicate adolescents using sedating antidepressants demonstrate a 2.8-fold higher incidence of prediabetes within two years [9, 34].
- Dependency Cycle: Rebound insomnia upon discontinuation creates a dependency cycle where 41% of adolescents resume medication to manage sleep disruption [28, 30], with discontinuation often driven by anxiety and agitation rather than traditional side effects [30].

These findings highlight a critical disconnect between clinical practice and evidence-based medicine. The Swedish Medical Products Agency explicitly states that doses ≤15 mg/day are not therapeutically effective for depression [1], yet our data show widespread prescribing at these subtherapeutic doses for sedation. Furthermore, the MIRAGE trial provides preliminary evidence for short-term mirtazapine use in older adults [4], but it does not address the specific risks for adolescents, whose developing brains are particularly vulnerable to chronic H1 receptor blockade [35, 39].

We call for urgent, evidence-based action:

1. Regulatory Alignment: Mirtazapine should be subject to the same stringent regulations as benzodiazepines, including mandatory prescriber education, limited initial prescription durations, and mandatory follow-up assessments to monitor for dependency and metabolic consequences [6, 25].
2. Prescribing Guidelines: Immediate revision of official dosage guidelines to reflect the evidence-based range of 5-30 mg/day, with explicit prohibition of off-label use for insomnia management in adolescents.
3. Expanded Non-Pharmacological Options: National initiative to expand access to Cognitive Behavioral Therapy for Insomnia (CBT-I), which remains available in fewer than 30% of Swedish primary care centers [8].
4. Metabolic Monitoring Protocol: Implementation of mandatory baseline and quarterly metabolic assessments (fasting glucose, lipid profile, BMI) for all patients receiving mirtazapine for more than 4 weeks.
5. Longitudinal Research: Prospective studies examining neurodevelopmental and metabolic outcomes in adolescents exposed to chronic H1 receptor blockade.

This trend represents more than a prescribing anomaly; it reflects a fundamental misalignment between clinical practice and adolescent health priorities. As mental health concerns rise among youth, we must ensure our interventions do not inadvertently create new health challenges. The time for policy action is now—before the current off-label prescribing patterns establish irreversible metabolic trajectories in a generation of Swedish adolescents.

### Study Limitations

#### This study has several limitations

First, prescription data reflects dispensed medications but not actual consumption or adherence. Second, while the registries do not capture indication data, our analysis of dosing patterns (8.84-10.76 mg/day) provides strong indirect evidence of off-label sedative use, as this range is subtherapeutic for depression but optimal for sedation [11, 33].

Third, we could not account for all potential confounding factors that might influence prescribing patterns, though the stark contrast with neighboring countries suggests policy rather than clinical factors drive the observed differences [5, 28].

Fourth, as a descriptive study, we cannot establish causality between policy changes and prescribing trends, though the temporal correlation with Sweden’s benzodiazepine restriction policies is compelling [6, 9].

Finally, while our dataset ends in 2017, the fundamental pharmacological properties of mirtazapine and the regulatory framework governing its use remain unchanged, suggesting the observed patterns likely persist.

## Data Availability

The dataset used in this study is publicly available at:

Kaggle Dataset:

https://www.kaggle.com/datasets/patricklford/antidepressant-usage-in-scandinavia-5-19yrs-old

Original sources:

- Sweden: https://sdb.socialstyrelsen.se
- Denmark: http://www.medstat.dk
- Norway: http://www.norpd.no

